# Milk antibody response after 3rd dose of COVID-19 mRNA vaccine and SARS-CoV-2 breakthrough infection and implications for infant protection

**DOI:** 10.1101/2022.12.12.22283367

**Authors:** Yarden Golan, Mikias Ilala, Caryl Gay, Soumya Hunagund, Christine Y. Lin, Arianna G. Cassidy, Unurzul Jigmeddagva, Lin Li, Nida Ozarslan, Ifeyinwa V. Asiodu, Nadav Ahituv, Valerie J. Flaherman, Stephanie L. Gaw, Mary Prahl

**Author notes:** <u>Co-corresponding authors:</u> Stephanie L. Gaw- Division of Maternal-Fetal Medicine, Department of Obstetrics, Gynecology, and Reproductive Sciences, University of California San Francisco, 513 Parnassus Ave, HSE16, Box 0556, San Francisco, CA 94143., Mary Prahl- Division of Pediatric Infectious Diseases and Global Health, Department of Pediatrics, University of California San Francisco, 550 16^th^ St. 4^th^ Floor. San Francisco, CA 94158. Authors contribution: YG, MP, and SLG designed the study. YG and MI conducted experiments. YG, acquired data, analyzed data, and was lead author of the manuscript. CG analyzed data, conducted statistical analysis assisted with editing the manuscript. SH and AC recruited participants, acquired data, and assisted with editing the manuscript. UJ, LL, NO, CYL collected samples and conducted experiments. IVA assisted with study and questionnaires design and editing and writing the manuscript. NA supervised the study and assisted in writing the manuscript. SLG and VJF designed and supervised the study and assisted in writing the manuscript. MP analyzed data, assisted with editing and writing the manuscript. YG, MP, SLG acquire funding for the study.

## Abstract

Anti-SARS-CoV-2 antibodies have been found in human-milk after COVID-19 infection and vaccination. However, little is known about their persistence in milk after booster vaccination and breakthrough infection. In this study, human-milk, saliva and blood samples were collected from 33 lactating individuals before and after mRNA-based vaccination and COVID-19 breakthrough infections. Antibody levels were measured using ELISA and symptoms were assessed using questionnaires. Evaluation of maternal and infant symptomatology revealed that infected mothers reported more symptoms than vaccinated mothers. We found that after vaccination, human-milk anti-SARS-CoV-2 antibodies persisted for up to 8 months. In addition, distinct patterns of human milk IgA and IgG production we observed after breakthrough infection compared to 3-dose vaccination series alone, indicating a differential central and mucosal immune profiles in hybrid compared with vaccine-induced immunity. To investigate passively-derived milk antibody protection in infants, we examined the persistence of these antibodies in infant saliva after breastfeeding. We found that IgA was more abundant in infant saliva compared to IgG and persist in infant saliva longer after feeding. Our results delineate the differences in milk antibody response to vaccination as compared to breakthrough infection and emphasize the importance of improving the secretion of IgA antibodies to human milk after vaccination to improve the protection of breastfeeding infants.

## Introduction

Exclusive breastfeeding is recommended for infants up to 6 months of age and is recommended by the American Academy of Pediatrics to be continued with the introduction of complementary foods to the infant diet for 2 years of age or longer [1]. Breastfeeding provides short and long-term protective effects from a number of diseases [1] and breastfeeding duration and exclusivity is specifically associated with reduced risk of lower respiratory tract infections in infants [2]. Human milk contains multiple factors that provide anti-viral protection to the infant including immune cells, extracellular vesicles, cytokines, enzymes and antibodies [3–5]. The breast is a unique organ in that despite not having a direct mucosal surface, it provides passive mucosal immunity including IgA, IgM, and IgG to the breastfeeding infant. IgA, the predominant human milk antibody, is typically present in its secretory form (sIgA) and provides passive mucosal defense for the infant’s respiratory and digestive systems [5–7]. In contrast, IgG, despite being the most prominent antibody in blood, is present in its monomeric form in human milk at lower levels than IgA or IgM, and helps provide protection against enteric pathogens [8,9]. Numerous studies have shown the presence of anti-SARS-CoV-2 antibodies in human milk after two doses of mRNA-based COVID-19 vaccines [10–21]. Specifically, IgA and IgG against the spike (S) protein of SARS-CoV-2 have been found in human milk after both vaccination and infection [7]. However, differential antibody dynamics based on the type of preceding antigen exposure — vaccination versus infection— has been described. Milk IgG increases significantly after the 2^nd^ vaccine dose, while secretory IgA significantly rises after SARS-CoV-2 infection with minimal increase of IgG [16,18]. As the COVID-19 pandemic and vaccine strategies have evolved over time, further information is needed on the potency and duration of the antibody response in milk beyond the 2^nd^ vaccine dose and the impact of hybrid immunity from breakthrough infections that have become increasingly common in the Omicron era.

Young infants are increased risk of severe disease and hospitalization from COVID-19 as compared to older children [22]. Current COVID-19 vaccinations are not approved until infants reach at least 6 months of age. Vaccination during pregnancy may provide some protection to the infant, as infants that were born to fully vaccinated mothers have a lower risk for COVID-19 infection[23] and hospitalization [24]. However, due to the lack of inclusion of lactating individuals in COVID-19 vaccination clinical trials, there is limited data on symptomatology and immune protection following vaccination and infection in lactating individuals and breastfeeding infants. Further information is needed on immune protection against COVID-19 during the vulnerable first months of infancy including the persistence of anti-SARS-CoV-2 antibodies in milk after vaccination and level of antibody transfer to the infant.

Here, we present longitudinal assessment of anti-SARS-CoV-2 milk antibody levels of lactating individuals after 2- or 3-dose vaccine series, as well as following breakthrough infection in vaccinated mothers. We assessed maternal and infant symptomatology after vaccination or infection. Lastly, we assessed the presence and duration of passively transferred antibodies in the saliva of breastfeeding infants.

## Results

### Participant cohort

Human milk samples were collected from 33 lactating individuals that received the first 2 doses of an mRNA-based COVID-19 vaccine (BTN162b2 or mRNA-1273) during pregnancy (n=25) or lactation (n=8) (**Table 1**). Twenty-six individuals from this cohort received the 3^rd^ dose of COVID-19 vaccine and reported their symptoms after vaccination (**Table 2**). Out of the 26 participants receiving 3^rd^ dose, 19 participants (3^rd^ dose subgroup) provided samples for antibodies assessment after 3^rd^ dose and their clinical characteristics are shown in **Table 3**. Of these 19 participants that received a 3^rd^ dose, 10 experienced a breakthrough infection from December 2021-March 2022, during the Omicron wave (SARS-CoV-2 B.1.1.529) in the San Francisco Bay Area (**Table 3**). Additional fourteen participants provided milk and/or saliva and infant saliva samples (after 2^nd^ or 3^rd^ dose).

**Table 1.**
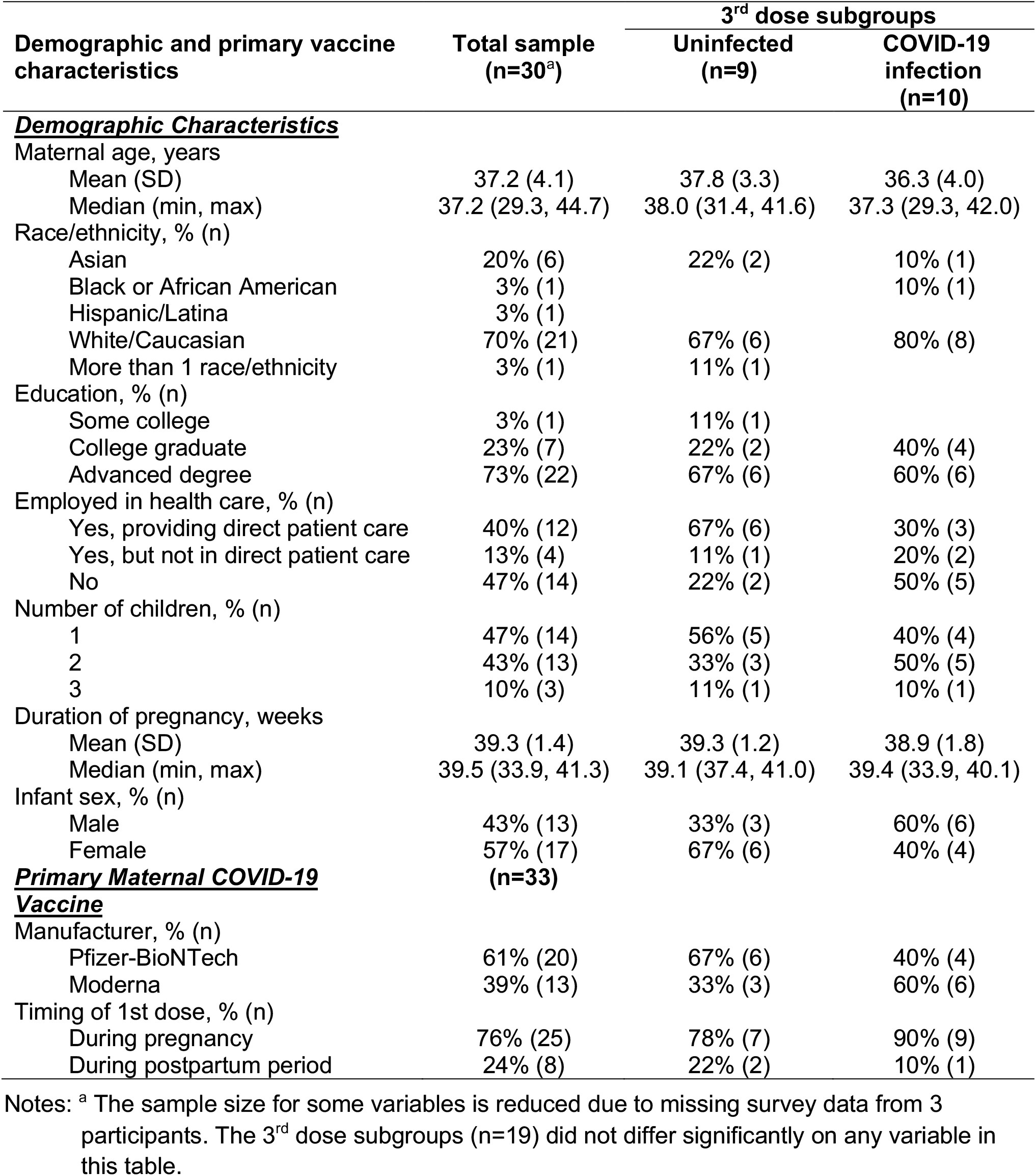
Sample characteristics, overall and for the 3^rd^ dose subgroups.

**Table 2.** Symptoms after 3rd vaccine dose.

| Symptoms | Total 3rd dose sample<br>(n=26) |
| --- | --- |
| Injection site symptoms, % (n) |  |
| Any injection site symptoms | 73% (19) |
| Pain | 62% (16) |
| Redness | 4% (1) |
| Swelling | 8% (2) |
| Itching | - |
| Rash near injection site | - |
| General symptoms, % (n) |  |
| Any general symptoms | 65% (17) |
| Fever | 12% (3) |
| Chills | 12% (3) |
| Headache | 27% (7) |
| Joint pain | 12% (3) |
| Muscle/body aches | 31% (8) |
| Fatigue or tiredness | 50% (13) |
| Lump/swelling in breast (opposite side of injection) | 4% (1) |

Note: Additional symptoms assessed but not reported by any participant after 3rd dose: nausea vomiting, diarrhea, abdominal pain, rash, mastitis, decreased milk supply or other symptom not listed above.

**Table 3.** Clinical characteristics of participants in the 3rd dose subgroups.

| Characteristics | 3rd dose subgroups |  |
| --- | --- | --- |
|  | Uninfected<br>(n=9) | COVID-19<br>infection<br>(n=10) |
| ***Maternal vaccine-related characteristics*** |  |  |
| Third vaccine dose received, % (n) |  |  |
| BTN162b2 (Pfizer-BioNTech) | 44% (4) | 30% (3) |
| mRNA-1273 (Moderna) | 56% (5) | 70% (7) |
| Infant age at 3rd dose, months |  |  |
| Mean (SD) | 6.8 (3.7) | 4.7 (3.4) |
| Median (min, max) | 5.5 (0.8, 11.9) | 5.2 (-1.7<sup>a</sup>, 9.4) |
| Days between third dose and symptom<br>assessment |  |  |
| Mean (SD) | 83 (92) | 121 (85) |
| Median (min, max) | 42 (15, 260) | 91.5 (29, 269) |
| ***SARS-CoV-2 infection characteristics<sup>b</sup>*** |  |  |
| COVID-19 infections, % (n) |  |  |
| Mother and infant |  | 80% (8) |
| Mother only |  | 20% (2) |
| Neither mother nor infant | 100% (9) |  |
| Infant age at time of maternal infection, months |  |  |
| Mean (SD) |  | 7.1 (3.5) |
| Median (min, max) |  | 7.8 (-0.8<sup>a</sup>, 12.0) |
| Days between maternal 3rd dose and<br>infection |  |  |
| Mean (SD) |  | 72 (38) |
| Median (min, max) |  | 72 (23, 141) |

Notes: The 3rd dose subgroups did not differ significantly on any variable in this table (comparisons exclude the 3 participants included in both subgroups).
<sup>a</sup> One infant had a negative age at both the maternal 3rd dose and maternal infection because both occurred during pregnancy, prior to the infant's birth.
<sup>b</sup> Of the 10 COVID-19 infections, 2 were diagnosed in late December 2021, 5 in January 2022, 2 in February 2022, and 1 in early March 2022.

### Symptomatology following 3^rd^ mRNA vaccine dose and/or COVID-19 infection

Patient reported symptoms were collected by REDCap surveys at least 2 weeks after exposure to a 3^rd^ mRNA vaccine dose and/or breakthrough infection. No severe symptoms were reported after the 3^rd^ vaccine dose in this cohort (**Table 2**). The most common maternal symptoms were pain in the injection site (reported by 62% of participants) or fatigue and tiredness (reported by 50% of participants). Maternal post 3^rd^ dose symptoms were significantly lower compared to symptoms reported in similar cohort of lactating individuals after 2^nd^ dose [12], and were not significantly different from reports after 1^st^ dose (**Figure 1**). In addition, symptoms reported in our cohort were similar to rates reported in larger cohorts [26]. When comparing post 3^rd^ dose and post-infection symptoms of the individuals with breakthrough infection in this cohort (n=10) we found that general symptoms were more likely to be reported by these participants after COVID-19 infection than after the 3^rd^ dose (p=0.025 for McNemar’s chi-square test) (**Table 4**). No infant symptoms were reported by mothers after receiving the 3rd dose (n=26); however, all infants that were infected with COVID-19 at the time of this study (n=8) had at least one symptom reported, including cough, runny nose, and fever (**Table 5**). No infants were hospitalized after COVID-19 infection in this cohort, but one infant required evaluation in the Emergency Department for their COVID-19 infection. Additionally, in seven of eight infected infants, surveyed mothers reported consultation with their physician about the infant’s COVID-19 infection (**Table 5**). The infected infants were on average 8 months old (range 5-12 months) and were not exclusively breastfed at this age (supplemented with formula or with complementary foods).

**Table 4.**
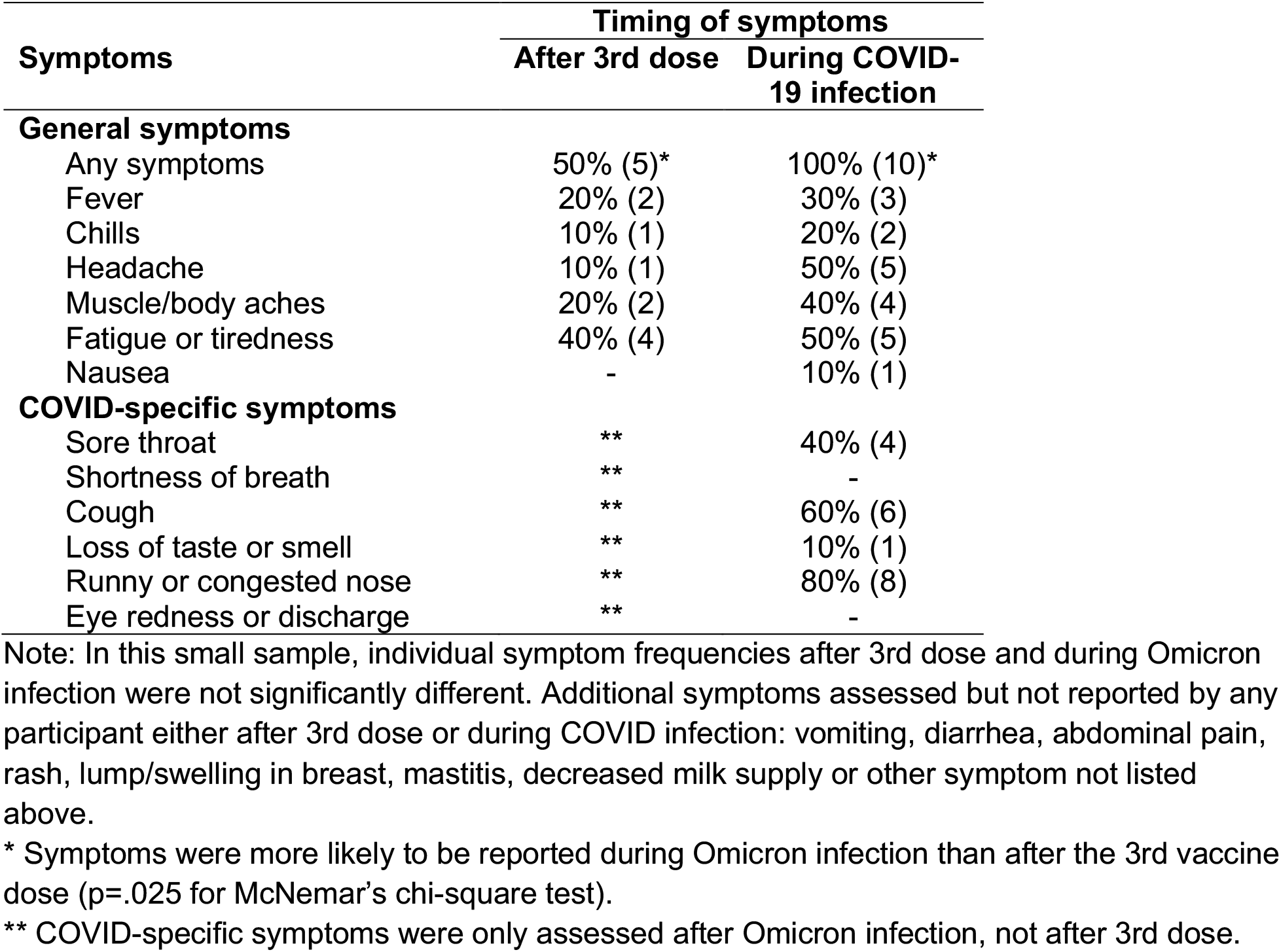
Comparison of post 3rd dose symptoms and COVID symptoms for participants with COVID-19 infection (n=10 paired analysis)

| Symptoms | Timing of symptoms |  |
| --- | --- | --- |
|  | After 3rd dose | During COVID-19 infection |
| **General symptoms** |  |  |
| Any symptoms | 50% (5)* | 100% (10)* |
| Fever | 20% (2) | 30% (3) |
| Chills | 10% (1) | 20% (2) |
| Headache | 10% (1) | 50% (5) |
| Muscle/body aches | 20% (2) | 40% (4) |
| Fatigue or tiredness | 40% (4) | 50% (5) |
| Nausea | - | 10% (1) |
| **COVID-specific symptoms** |  |  |
| Sore throat | ** | 40% (4) |
| Shortness of breath | ** | - |
| Cough | ** | 60% (6) |
| Loss of taste or smell | ** | 10% (1) |
| Runny or congested nose | ** | 80% (8) |
| Eye redness or discharge | ** | - |

Note: In this small sample, individual symptom frequencies after 3rd dose and during Omicron infection were not significantly different. Additional symptoms assessed but not reported by any participant either after 3rd dose or during COVID infection: vomiting, diarrhea, abdominal pain, rash, lump/swelling in breast, mastitis, decreased milk supply or other symptom not listed above.
\* Symptoms were more likely to be reported during Omicron infection than after the 3rd vaccine dose (p=.025 for McNemar's chi-square test).
\*\* COVID-specific symptoms were only assessed after Omicron infection, not after 3rd dose.

**Table 5.** Infant symptoms after 3rd maternal vaccine dose and during COVID-19 infection.

| **INFANT SYMPTOMS AFTER THIRD MATERNAL VACCINE DOSE** | **% (n)** |
| --- | --- |
| None reported among the 26 participants who completed the symptom survey after their third vaccine dose | 0% (0) |
| **INFANT SYMPTOMS DURING COVID-19 INFECTION (n=8)** | **% (n)** |
| Any symptoms | 100% (8) |
| Cough | 88% (7) |
| Runny nose | 88% (7) |
| Fever | 50% (4) |
| Fatigue | 25% (2) |
| Loss of appetite | 38% (3) |
| Eye redness | 12% (1) |
| Diarrhea | 12% (1) |
| Swollen lymph nodes | 12% (1) |
| Joint pain | 12% (1) |
| Muscle/body aches | 12% (1) |
| Other symptoms: | 25% (2) |
| Mildly fussy/woke up more frequently during the night. Mild fever and fussiness/fatigue lasted ~24 hours but runny nose persisted for a week |  |
| Mild increased work of breathing and mild tachypnea |  |
| **TREATMENT FOR INFANT COVID INFECTION (n=8)** |  |
| Consulted with pediatrician regarding COVID-19 infection | 88% (7) |
| Treated with anti-pyretics | 12% (1) |
| Evaluated in Emergency Department for COVID-19 infection | 12% (1) |
| Infant was hospitalized for COVID-19 Infection | - |

Symptoms asked about, but not reported for any infant: rash, hand or foot swelling, redness of tongue, shortness of breath, chest discomfort/pain, loss of taste or smell, headache, dizziness, vertigo, insomnia, hair loss, persistent sweating, impaired memory, poor concentration.

**Figure 1:**
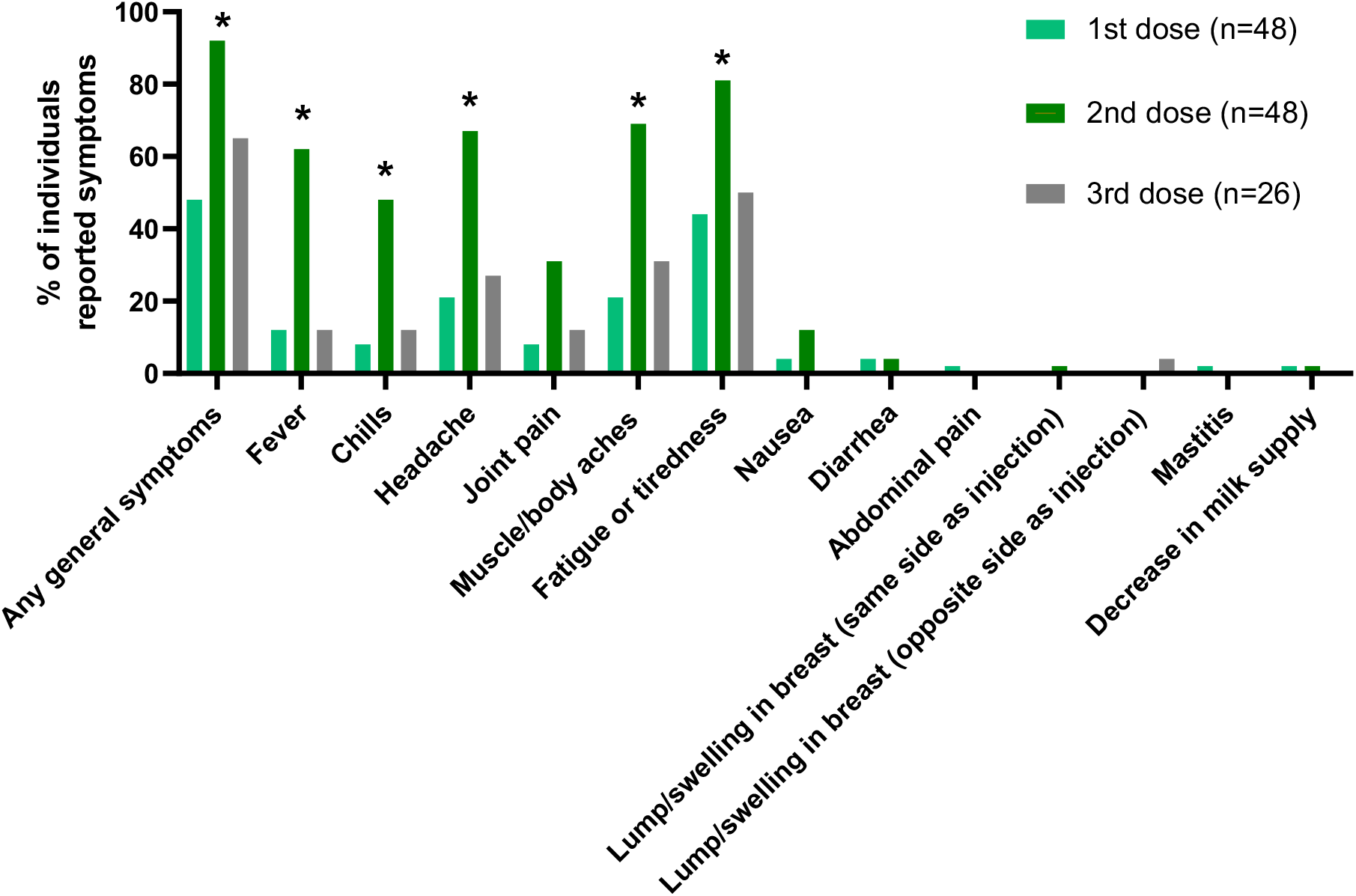
Comparison of symptoms reported by participants after 1^st^, 2^nd^ and 3^rd^ mRNA-based vaccine. Fisher’s Exact test for independent samples was perform to compare symptoms reported after 3^rd^ dose to those reported after 1^st^ and 2^nd^ dose in lactating individuals, as previously reported [12]. Asterisks indicate symptoms significantly different between 2^nd^ and 3^rd^ dose (p-value <0.01). No significant differences were observed between 1^st^ and 3^rd^ dose.

### Longitudinal persistence of anti-SARS-CoV-2 milk antibodies after vaccination and differential milk IgA responses following breakthrough COVID-19 infection compared to post-vaccination

Milk anti-Spike antibodies were detected 6-8 months following the 2^nd^ dose, but milk Spike IgG significantly decreased over time — with only 52% (10 of 19) of individuals had detectable antibodies in milk prior to 3^rd^ dose boost vaccination. (**Figure 2A**). In contrast to IgG, 16 of 19 (84%) of individuals maintained detectable levels of milk anti-Spike IgA after the 2^nd^ dose, and prior to 3^rd^ dose boosting, but there was also a significant decrease in these antibody levels over time (**Figure 2B**). After the 3^rd^ dose, milk anti-Spike IgG levels increased significantly, and were significantly higher compared to their levels following the 2^nd^ dose (**Figure 2A**). Milk anti-Spike IgA levels also trended higher after receipt of the 3^rd^ dose but was not statistically significantly increased over pre-boost levels and their levels were similar to the post 2^nd^ dose timepoint indicating a persistence of anti-Spike milk IgA over time after primary vaccination series, but a lack of significant boosting of milk anti-Spike IgA levels after the 3^rd^ dose. Both IgG and IgA levels decreased 5 months after the 3^rd^ dose, but in contrast to the pre-boost timepoint all participants had detectable IgG levels and only 3 of 5 (60%) had detectable IgA levels at this time point (**Figure 2A and 2B**). Individuals with breakthrough SARS-CoV-2 infection after their 3^rd^ dose had significantly higher levels of IgA in their milk following infection (**Figure 2B**) compared to individuals after 2^nd^ and 3^rd^ vaccine doses. However, milk anti-Spike IgG levels after infection were comparable to the levels after 3^rd^ dose (**Figure 2A**). Furthermore, we found higher levels of anti-Spike IgA antibodies in the plasma of lactating individuals after infection compared to after the 3^rd^ dose (**Figure 2C**). There was also a positive correlation between anti-Spike IgA antibodies levels in milk and maternal plasma after the 3^rd^ dose (r= 0.8091 and p=0.0039 **Figure 2D**), and the correlation between milk and blood IgA levels was even stronger after infection (r=0.937 and p-value 0.0006).

**Figure 2:**
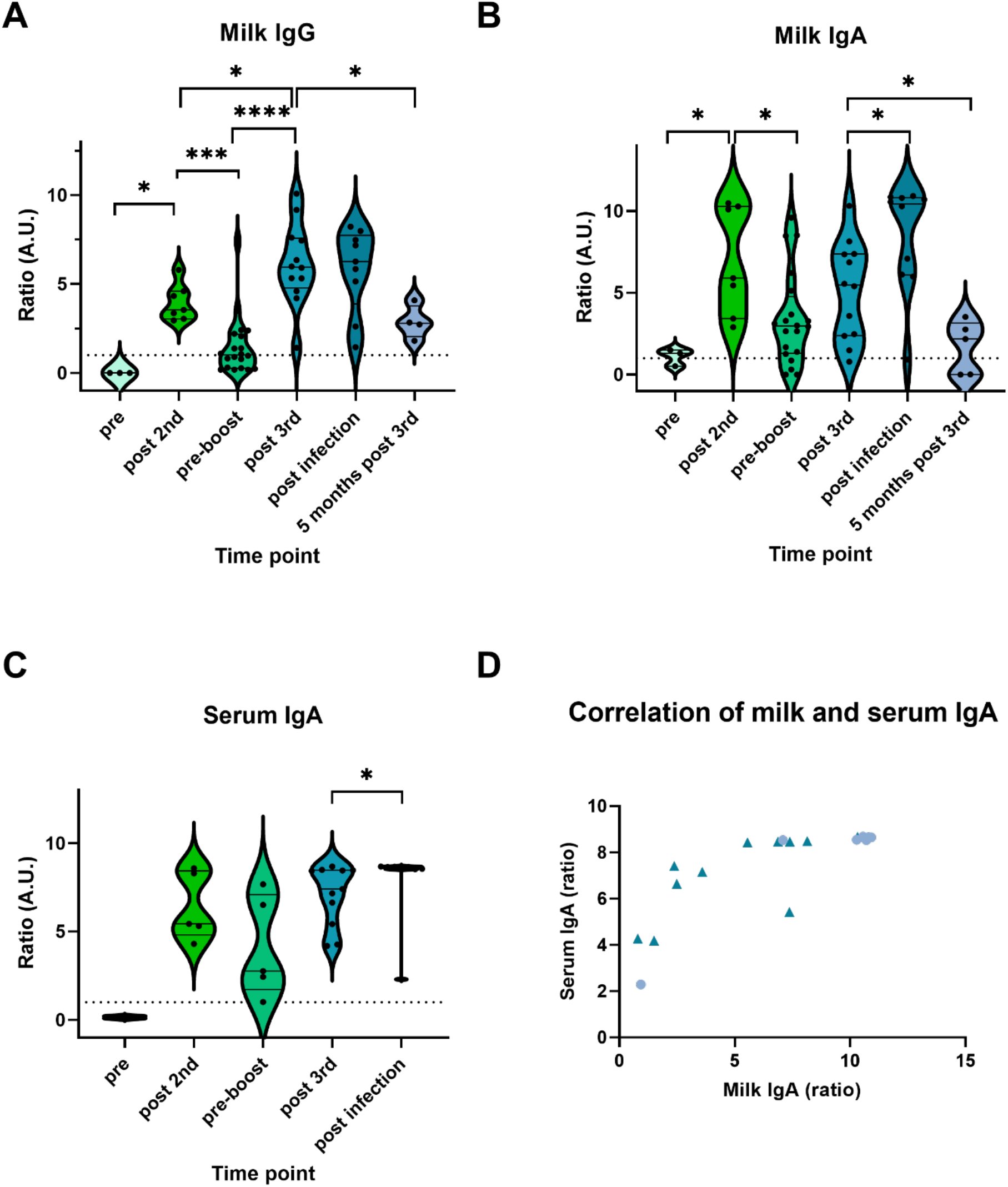
Anti-Spike antibodies response to vaccination and infection. Anti-Spike IgG (A) and IgA (B) were measured in human milk samples and IgA was measured in serum samples (C) by ELISA, at multiple time points as represented in the X axis. Samples collected before any COVID-19 vaccine administration (pre), 41±8 days after the 2^nd^ dose (post 2^nd^), 223±36 days after the 2nd dose (pre-boost), 42±15 days post 3rd dose (post 3rd) and 142±10 days post the 3rd dose (long post 3rd) and 36±15 days post infection with SARS-CoV-2 (post infection). Dotted lines indicate the lower cut-off (above 1 considered positive). Upper limit of detection was at a ratio of 8.5. Asterisks represent p-values: *= p-value <0.05, **= p-value <0.01, ***= <0.001, ****= <0.0001 as determined by unpaired Mann-Whitney test. D) Two-tailed Spearman correlation was used to correlate milk and serum anti-Spike IgA levels at Post-Dose 3 (triangles) and Post infection (circles).

### Persistence of maternal milk-derived SARS-CoV-2 antibodies in infant saliva after breastfeeding

Milk antibodies may provide protection to the infant at the oropharyngeal mucosal surfaces, but little is known regarding the stability of these antibodies in the infant mouth after breastfeeding. To answer this question, we investigated the stability and persistence of milk antibodies in infant saliva after breastfeeding using saliva samples collected from infants at multiple time points after feeding. We compared these to antibody levels in maternal milk and saliva samples collected the same day as the infant. We found a positive correlation between anti-Spike IgA levels in milk and maternal saliva samples (**Figure 3A)**, as well as a positive but non-significant correlation for milk and maternal saliva anti-Spike IgG antibodies (**Figure 3B**). We next evaluated infant saliva samples collected after feeding by mothers who had detectable anti-Spike IgG or IgA in their milk. Anti-Spike IgA levels were found to be significantly higher in infant saliva over time after feeding compared to IgG antibodies, with 6/11 (55%) infants having detectable antibodies immediately after breastfeeding and 3/11 (27%) infants remaining positive at all time points until the next feeding. We found that IgG antibodies were less abundant in the infant’s saliva after feeding, with all except one infant’s samples below the assay cut-off **(Figure 3C and 3D)**. Of note, we could not identify any correlation between infant age or lactation exclusivity to infant saliva antibodies, which might be due to the small sample size of this study.

**Figure 3:**
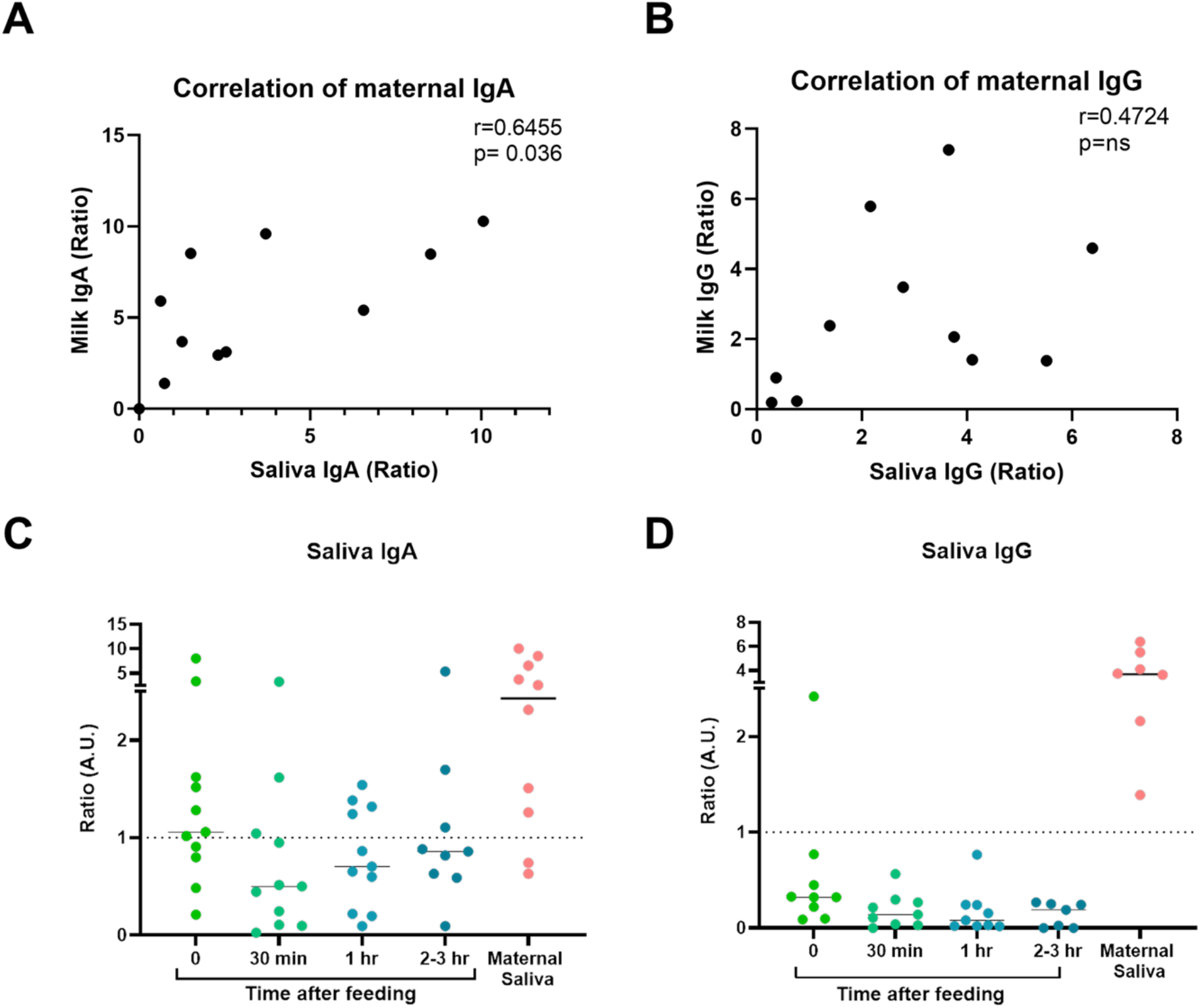
Maternal saliva antibodies response and presence of milk antibodies in infant saliva after feeding. Two-tailed Spearman correlation was used to correlate milk and maternal saliva anti-Spike IgA levels (A) and IgG (B). In addition, anti-Spike IgA levels (C) and IgG (D) infant saliva samples were measured at multiple time points, immediately after breastfeeding (0), 30 minutes (30 min) after feeding, 1 hour after feeding (1 hr) and before next feeding (2-3 hours after feeding). Maternal saliva was collected at the same day for comparison. Dotted lines indicate the lower cut-off (above 1 considered positive).

## Discussion

We found that mRNA-based vaccines administered in pregnancy or during lactation stimulated increased anti-SARS-CoV-2 antibody (specifically anti-Spike antibody) levels in human milk that persisted for up to 8 months after vaccination. In addition, we found that a 3^rd^ dose (booster) significantly increased the IgG antibody levels in milk. These findings highlight the importance of vaccine boosting during pregnancy and lactation in providing additional transfer of antibodies through human milk, which may provide protection against SARS-CoV-2 to young infants, who are currently not eligible for COVID-19 vaccination until they reach 6 months of age.

In addition, we performed a comparative analysis of anti-Spike milk IgG and IgA levels that are induced after 2 doses vs 3 doses of the mRNA vaccine, as well as SARS-CoV-2 breakthrough infection. Although anti-SARS-CoV-2 IgA antibodies are induced after vaccination, their levels are significantly boosted after natural infection as compared to vaccination alone. In contrast, milk anti-SARS-CoV-2 IgG levels did not significantly increase after breakthrough infection. Of note, milk IgG levels were very high after the 3^rd^ dose and may have already reached peak levels at the time of breakthrough infections, which occurred within 7 weeks of the 3^rd^ dose in our cohort. Previous studies have also shown similar patterns of IgA in milk and saliva [27,28], indicating that exposure to SARS-CoV-2 infection has a greater effect on mucosal IgA secretion compared to vaccination. Interestingly, mucosal and systemically-delivered influenza vaccines result in similar increases in influenza specific IgA antibodies in milk [29] suggesting that factors in addition to exposure location may affect the production of milk IgA antibodies.

IgA antibodies play a critical role in humoral immune response and virus neutralization, with peripheral expansion of IgA plasmablasts found in SARS-CoV-2 infected patients shortly after the onset of symptoms [30]. Our results demonstrate a positive correlation between maternal IgA levels in milk and plasma, as well as milk and saliva. Previous studies have shown that mucosal immunity in the bronchoalveolar lavage fluid is weaker after vaccination compared to post-infection immunity [31]. In addition, COVID-19 infection prior to vaccination was shown to induce a better secretion of antigen-specific mucosal secretory IgA to the saliva, compared to vaccination alone [32]. Our findings further suggest that mRNA-vaccines induced an IgA response in milk and in blood, but to a lower extent compared to hybrid immunity from vaccination and COVID-19 infection. To the best of our knowledge, our work is the first to compare boosting of milk antibody levels after the 3rd dose versus infection during the time that the Omicron variant was the predominant circulating strain.

In addition, we found that IgA antibodies are more abundant in infant saliva at multiple time points after breastfeeding compared to IgG; therefore, developing vaccines that improve the secretion of IgA antibodies to milk (and other mucosal organs) might also better contribute to infant (and mother) protection against infection. Larger studies are needed to evaluate the protective effects of anti-SARS-CoV-2 milk-derived antibodies on breastfed infants.

Eight infants in our cohort were infected with SARS-CoV-2 during the study period (when the Omicron variant was predominant [33]), in the setting of maternal infection, despite their mother having received the 3^rd^ vaccine dose. Regardless of the persistence of anti-SARS-CoV-2 antibodies in milk over time, passively derived milk antibodies alone were insufficient to fully protect against infection, possibly due to immune evasion from vaccine-induced antibodies by the Omicron variant, and/or weaker protection provided by milk antibodies compared to passively-acquired transplacentally transferred IgG systemic antibodies that wane after birth [34,35]. All infants infected in this cohort were older then 5 months, so their transplacental antibodies were lower or absent at time of infection, and most of them were not exclusively breastfed when infected (were supplemented with baby formula or complementary foods). However, due to our limited sample size, we were unable to assess the level of protection provided by transfer of vaccine-related antibodies in human milk, as compared to infants with no SARS-CoV-2 vaccine-related milk antibodies. The Center for Disease Control and Prevention (CDC) reported that during the early 2022 Omicron variant peak, infants hospitalization rates were 5 times higher compared to during the Delta variant peak [22]. All COVID-19 infected infants in this cohort presented with symptoms, and one infant was admitted for evaluation in emergency care unit for COVID-19 symptoms. These results underscore the importance of both passive maternally-derived and early infancy vaccination protection for this vulnerable infant population. In contrast to infant infected with SARS-CoV-2, no infants in our cohort were reported to have symptoms following maternal COVID-19 vaccination during lactation. In a larger cohort which examined 10,278 participants after 3rd dose, 1.2% of mothers reported any issue in their infant after vaccination during lactation [26]. These reports emphasize the importance of including lactating individuals in clinical trials, to be able to examine the direct effect of vaccine administration on infant symptoms, in compared to placebo group, which are currently absent.

In summary, we found that human milk antibody levels increase significantly after the 2^nd^ vaccine dose and can maintain high levels in milk up to 8 months post vaccination in some individuals. Boosting with a 3^rd^ vaccine dose significantly increases IgG antibody levels that remain elevated for at least an additional 5 months post-booster vaccination in milk. Milk IgA antibodies were much more significantly increased after breakthrough SARS-CoV-2 infection, compared to vaccination alone. It is specifically important as based on our results, IgA as compared to IgG antibodies were more stable in the infant mouth after feeding — and may be more important in infant protection against SARS-CoV-2 infection. Further large-scale cohort studies of vaccinated lactating individuals are needed to better understand the role of milk antibodies in infant protection from COVID-19 infection. Future vaccine development should focus on the induction of milk IgA antibodies to enhance infant protection during lactation.

## Methods

### Participant cohort and data collection

The institutional review board of the University of California, San Francisco, approved the study (#21-33621). Written informed consent was obtained from all study volunteers as part of the COVID-19 Vaccine in Pregnancy and Lactation (COVIPAL) cohort study. Pregnant or lactating mRNA COVID-19 vaccination recipients were enrolled from December 2020 to April 2022. Clinical data and symptoms were collected by medical record review and REDCap questionnaires and characterized in **Table 1**. Participants were surveyed following each COVID-19 vaccine dose, which included questions about maternal and infant symptomology after maternal 3^rd^ dose. In February 2022 all COVIPAL participants were surveyed if they had a new diagnosis of SARS-CoV-2 infection since the last vaccine dose and if the participant was willing to provide post-infection biospecimen samples. Individuals with SARS-CoV-2 breakthrough infection (confirmed by PCR or rapid antigen testing) were administered questionnaires to capture maternal and infant post infection symptoms. For individuals at the time of post infection survey that had not yet completed their 3^rd^ dose questionnaire, their 3^rd^ dose symptoms were captured at the same time of their post infection symptoms (up to 6 months after receiving the 3^rd^ vaccine dose). No infants were vaccinated during the study period.

### Milk sample collection and processing

Milk samples were collected at the following time points: 1) Pre-vaccine 2) Post-Dose 2 (range 4.7 to 7 weeks following 2^nd^ dose; 3) Pre-Boost (prior to 3^rd^ dose, range 26-37 weeks following 2^nd^ dose); 4) Post-Dose 3 (range 4-10 weeks following 3^rd^ dose; 5) 5 months after Post-Dose 3 (range 18-21 weeks after 3^rd^ dose) and 6) Post-Infection (range 2-7 weeks following breakthrough infection accruing after 3-dose vaccination series). Fresh human milk samples were self-collected by participants into sterile containers. Milk samples were either processed immediately by the study staff or frozen by mothers in their home freezer as soon as possible after pumping. Samples were transported on ice from participant’s home to the lab for processing. Milk was aliquoted and stored at −80°C until analyzed.

### Blood sample collection and processing

Paired maternal blood samples were collected at the same time points as described above for milk samples from a subset of participants (n=18). Whole blood was collected into EDTA tubes. Plasma was isolated from whole blood by centrifugation and immediately cryopreserved at −80°C until analysis.

### Infant and maternal saliva sample collection

To evaluate the duration of persistence of antibodies in the infant’s mouth after breastfeeding, we collected saliva samples from breastfeeding infants at the following timepoints: 1) immediately after breastfeeding 2) 30 min after breastfeeding 3) 60 min after breastfeeding and 4) before next breastfeeding (2-3 hours after feeding). Paired maternal saliva and milk samples were collected from the infant’s mother on the day of infant collection (n=11). Maternal saliva samples were collected with OraSure collection kits by placing the swab in the mouth for 5 minutes until saturated. For infant saliva samples, sponges for assisted saliva collection were used and were inserted to OraSure collection tubes (Product CS-2, DNAGenotek Inc, Ontario, Canada).

### ELISA assay

Anti-Spike ELISA assay (Euroimmune, Germany) was used to measure IgA and/or IgG levels in blood, skim milk, and saliva samples. Plasma and skim milk samples were thawed on ice. After thawing, milk fat was separated by cold centrifugation (10,000g for 10 min, 4°C) and diluted 1:4 with the provided diluent buffer, and examined using the manufacturer’s protocol as described, with an additional blocking step with 5% BSA in TBS with 0.5% Tween 20 for 30 min before loading the samples as recommended to increase specificity [25]. Plasma samples were diluted 1:101 and were examined by the same protocol as milk samples. OD values of samples were calculated by dividing by the calibrator OD value (provided with the kit); values with sample:calibrator ratio greater than 1 were considered positive. Saliva samples were concentrated using Amicon Ultra Centrifugal Filters (UFC5199BK, Millipore Sigma, USA), as previously described [25]. Milk samples were analyzed in duplicate; and saliva samples were analyzed once for each antibody (IgA and IgG) due to limited sample availability.

### Statistical analysis

Spearman analysis was used for correlation between milk and maternal saliva antibodies levels. Mann-Whitney test was used to evaluate differences between the different time points in milk and plasma antibody levels. Wilcoxon matched-pairs signed rank test was used to compared IgA and IgG levels in infant saliva at the different time points. Comparison of symptoms reported by each participant after 3^rd^ dose and after infection was performed by McNemar’s chi-square test. Fisher’s Exact test was used to compared post vaccine symptoms after 1, 2 or 3 doses. Data about 1^st^ and 2^nd^ dose were previously reported [12]. Statistical analysis was performed using Prism version 9.1.0 (GraphPad) and Stata Version 15.0 were used for analyses.

## Data Availability

All data produced in the present study are available upon reasonable request to the authors

